# Effects of diet, activities, environmental exposures and trimethylamine metabolism on alveolar breath compounds: protocol for a retrospective case-cohort observational study

**DOI:** 10.1101/2021.01.25.21250101

**Authors:** Irene S. Gabashvili

## Abstract

**Background:** Exhaled breath contains thousands of volatile organic compounds (VOCs) that reflect on biochemical and biophysical activities both outside and within the human body. Breath analysis could provide non-invasive, cost-effective, real time early disease diagnosis and monitoring of therapeutic responses.

**Objectives:** The primary objective of this study was to assess the effectiveness of alveolar breath testing in diagnosing idiopathic systemic body and breath odors. Key secondary objectives were to assess if breath tests can reliably differentiate subtypes of idiopathic malodor in different environments and dietary regimens, and to map metabolites to biomedical functions and pathways.

**Study Design:** The basic design was to measure a cohort of idiopathic odor in order to identify potential molecular correlates with genotypic and phenotypic variables. Participants were subdivided in several different ways allowing for different cases and controls within the cohort, using prior and later test results and observations. Thus, this study was an observational retrospective case-cohort/nested case-control study.

**Setting/Participants:** Participants were recruited online via MEBO and TMAU support groups and on site, during the 3rd Annual MEBO Research conference held at Miami South Beach on June 23, 2012 and local meetups of support groups (Miami, Florida; New York, New York; Chicago, Illinois, US and Birmingham, England). Study population is individuals self-reporting systemic idiopathic malodor production. Inclusion criteria were good general health, desire and ability to travel to one of the participating sites and pay the lab fee. Exclusion Criteria were medical conditions that could prevent participation and age under 18.

**Study Interventions and Measures:** The main study procedure was the application of a rapid point-of-care breath testing system to collect and concentrate alveolar breath VOCs on a sorbent trap, using breath collection apparatus (BCA) 5.0. Samples were sent to central laboratory and analyzed with gas chromatography and mass spectroscopy. In addition, the participants filled out food frequency questionnaires and were offered to use [Aurametrix](https://aurametrix.com/), online software tool based on a participant-initiated ecological momentary assessment approach, allowing to recall the events at any time later. The tool analyzed dietary intakes, activities and environmental exposures for both individual and aggregate level data.

The primary endpoint was the composition of VOCs in breath samples, while diet and activity data, and results of alternative testing assessments were secondary endpoints. The main study outcome measure is the diagnostic accuracy of alveolar breath test in differentiating profiles of two main pre-defined sub-cohorts. Index of concordance with accuracy, sensitivity, specificity, positive predictive value and negative predictive value will be reported. A number of factors was assessed for confounding.

## Background Information and Rationale

### Introduction

Measurement of volatile organic compounds (VOCs) in the exhaled breath has demonstrated early proof of concept in the diagnosis of metabolic, immune, gastrointestinal, neurological and psychiatric disorders, cardiovascular, respiratory, urinary, inflammatory and infection diseases, and in differentiating subtypes of multiple cancers [1-28].

Since the only symptom of idiopathic malodor is an unpleasant smell with intensity exceeding socially acceptable levels, it’s logical to assume that volatile compounds responsible for it can be detected in the air surrounding the body, and hence in the exhaled breath, and provide information about underlying causes.

### Relevant Literature and Data

Idiopathic malodor – body and breath odor arising spontaneously without any known pathology or cause – is often assumed to be an undiagnosed case of trimethylaminuria (TMAU) [29]. TMAU is a metabolic disorder causing elevated levels of a foul-smelling tertiary amine trimethylamine (TMA) not oxidized into odorless trimethylamine N-oxide (TMAO), leading to TMA/TMAO ratio in urine exceeding 5% [30,31].

TMAU can be difficult to diagnose without a choline challenge test [32,33], because metabolite production fluctuates, depending on diet and other activities of daily living. Metabolite analysis focused on detection of trimethylamine (TMA) and trimethylamine N-oxide (TMAO) so far has not been very successful. Even the most obvious genetically diagnosed cases are not always confirmed metabolically [34]. No comprehensive analysis for non-targeted screening by gas chromatography-mass spectrometry have been yet, to our knowledge, reported for exhaled breath in TMAU and TMAU-like conditions.

Idiopathic malodor can be also associated with an understudied and hard-to-diagnose psychiatric condition Olfactory Reference Syndrome (ORS) (subtype of a Body Dysmorphic Disorder) and a similar Japanese-culture-bound condition “taijinkyofusho” [34]. Olfactory reference syndrome is a nonorganic psychiatric disorder in which the chief complaint is the patient’s awareness of his body odor that surrounding people do not detect [35]. ORS may be misdiagnosed as schizophrenia [36] found to have a distinct VOC signature in exhaled breath [37].

Our recent work [38] showed that non-syndromic systemic malodor produces a distinctive cutaneous microbial signature for different subtypes of TMAU as well as TMAU-negative metabolic body odor (MEBO) and “People are allergic to me” (PATM) conditions. Our earlier studies identified potential metabolic biomarkers of MEBO and PATM in blood [39], urine [40] and gastrointestinal tract [39], suggesting that volatile organic compounds can potentially be used as diagnostic signatures to distinguish between sub-cohorts of idiopathic malodor.

Exhaled breath analysis is one of the most promising non-invasive diagnostic techniques. Proposed exploratory study will compile prior knowledge and molecular information for each alveolar metabolite from the literature and online databases to ensure the selection of the most reliable candidates.

Notable published information includes volatiles pertaining to metabolic pathways of interest (41), volatiles produced by anaerobic Gram-negative bacteria or oxidative pathways identified in halitosis [42-45], and other volatiles released into the air by bacteria [46], viruses [47] and cell lines growing on defined media [48]

### Compliance Statement

This study will be conducted in full accordance all applicable Federal and state laws and regulations including 45 CFR 46, 21 CFR Parts 50, 54, 56, and the Good Clinical Practice: Consolidated Guideline approved by the International Conference on Harmonisation (ICH).

Both original and secondary studies were reviewed by MEBO Research Institutional Review Board registered with the Office of Human Research Protections on February 3 2012 (IRB00008681, FWA00018466). Current FWA00026547 (IRB00010169) expires on 02/14/2023.

Protocol Number for the primary prospective study: MR 2012-01

Date of IRB Approval: February 28, 2012

Protocol Number for the retrospective secondary study: 20111001005MEBO

Date of IRB Approval: February 15, 2018

All episodes of noncompliance will be documented.

The investigators will perform the study in accordance with this protocol, will obtain consent, and will report unanticipated problems involving risks to subjects or others in accordance with all federal requirements. Collection, recording, and reporting of data will be accurate and will ensure the privacy, health, and welfare of research subjects during and after the study.

## Study Objectives

The purpose of the study is to determine the relationship between volatile organic compounds in exhaled breath and subtypes of systemic malodor conditions.

### Primary Objective

The primary objective of this study is to establish the accuracy of the breath test for determining the presence of systemic body odor. All subjects were asked to provide a single sample of breath, report their symptoms, medical history and allow the study team to collect information about their other diagnoses.

### Secondary Objectives

The secondary objectives are to determine if there is a relationship between volatile organic compounds and results of other biomedical measurements and diagnostic tests.

## Investigational plan

The original prospective study was designed as a case-control, comparing individuals reporting systemic malodor with a control population. This study employs alternative cost-effective sub-cohort sampling strategies to conduct biomarker research such as nested case–control and other variations of case-cohort design. Lag interval information regarding volatiles in exhaled breath vs results of other measurements will be collected retrospectively.

### General Schema of Study Design

This study focusing on composition of metabolites in exhaled breath has a retrospective observational design. Subjects (n=23) were recruited from cohorts of prior MEBO Research studies’ and support groups. Potential subjects were screened using the protocol inclusion and exclusion criteria. To collect medical data for screening purposes, an in-person consent was obtained from each participant. Information regarding exposure and events that occurred between trial end (n=22 had provided processable breath samples) and registry enrollment will be collected retrospectively. All subjects will be included in the database.

### Study Duration, Enrollment and Number of Sites

Case reports will be included in the study if breath samples and daily logs were collected between 10/1/2011 and 12/31/2013. Follow-up information though 12/12/2020 will be also included, as well as medical history preceding the initial breath sampling. Study duration will be from study initiation or subject enrollment and registration and until the subject withdraws or is withdrawn or discontinued from the study.

The study sites were in Miami, Florida; New York, New York; Chicago, Illinois in the USA; London and Birmingham in England, UK.

### Study Population

In order to be eligible, participants must fulfil the following criteria:

#### Inclusion Criteria

1. Adults, aged 18 years or older.
2. Good general health
3. Willing and able to travel to one of the participating sites

#### Exclusion Criteria

1. Medical conditions that, in the opinion of the investigator, would prevent participation
2. elect not to participate in the study

Subjects that do not meet all of the enrollment criteria may not be enrolled. Any violations of these criteria will be reported in accordance with IRB Policies and Procedures.

## Study Procedures

### Screening, Observations and Measures

This study consists of one in-person data collection time point. Procedures to be performed at this visit include informed consent, medical records review and breath sample collection, as shown below. The figure (modified from [49]) shows BreathLink^™^, a breath collection apparatus for gathering, condensing and analyzing VOCs in human breath [50]. In the upper inset (labeled 1) of this Figure, an individual wears a nose-clip and respires for two minutes through a disposable-valved mouthpiece unit and a bacterial filter. Inset in the lower right corner (labeled 2) shows a sealed sorbent trap. VOCs in alveolar breath and room air are collected on to separate sorbent traps which are hermetically sealed for shipping to the laboratory. Lower left inset (3) is an example gas chromatogram of breath. Breath samples are desorbed onto an automated devise for thermal desorbtion that concentrates the VOCs almost 1 million-fold prior to analysis by gas chromatography and mass spectroscopy. A chromatogram of breath typically contains 150–200 peaks, each eluting with a different retention time. Each peak usually, but not invariably, represents one VOC. The inset at the upper left-hand side (4) shows a typical mass spectrum retrieved for one chromatogram peak. The probable chemical structure of the VOC is inferred from its resemblance to another mass spectrum in the computerized library [51]. In this example, Undecane (CAS# 1120-21-4), was determined to be the most likely correct. In order to identify candidate biomarkers of a medical condition, after primary identification of VOCs, mass spectra are analyzed for secondary identification. BreathLink^™^ has been used to diagnose patients with active pulmonary tuberculosis [52], breast cancer [53,54] and lung cancer [55].

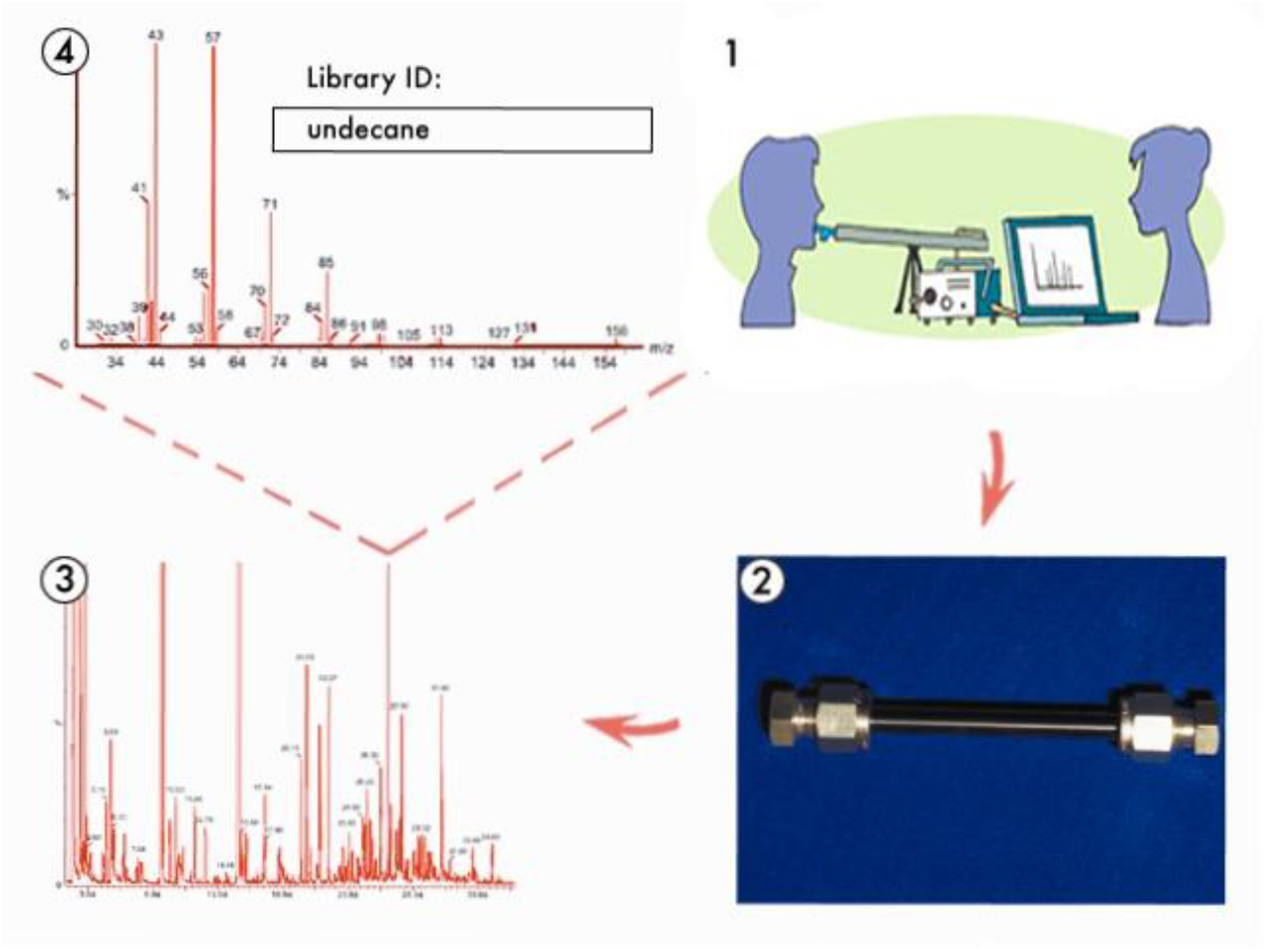

### Observational Period

Since several studies and follow-ups were done before and after the end of prospective breath testing trial in December 2013, results of that studies will be included in retrospective data analysis.

#### Biolab Study

Biolab study (2009-2012) was designed as a prospective cohort study to evaluate the potential of diagnostic procedures in defining populations of patients with symptoms of systemic body odor and/or halitosis [39]. It was registered on clinicaltrials.gov as NCT02692495

#### MEBO TMAU Test Program

Measurement of Trimethylamine and Trimethylamine N-Oxide (TMA :TMAO) ratio in urine (2012-2017) was performed in hundreds of patients around the world, including some of the participants of this alveolar breath testing study.

#### Aurametrix Study

Aurametrix software solution was designed to help people understand relationships between their diets, medications, environmental exposures, daily activities and symptoms such as systemic malodor [56,57]. It was available to the participants of this trail from October 2011 till the end of 2013. Aurametrix platform was used as a study diary as well as a research tool providing feedback to improve users’ wellbeing.

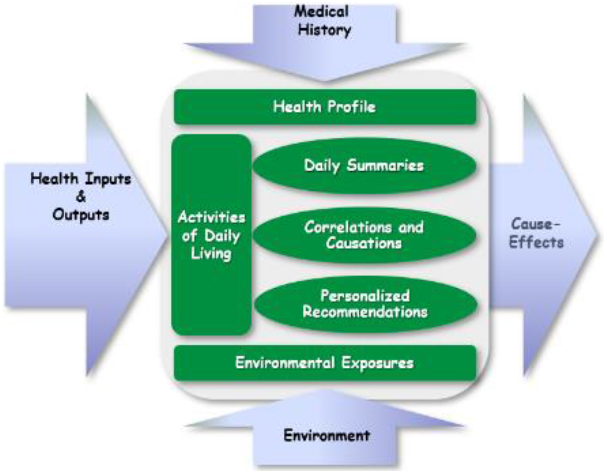

The system is based on a rich, multi-domain knowledgebase, food and symptom ontologies and automated analysis of existing health conditions, medications and supplements, diet, metabolic phenotypes, exercise, local air quality, allergen levels, and many other determinants of health.

#### Urine Metabolome Study

Exploratory Study of Relationships Between Malodor and Urine Metabolomics (2016-2018) was designed to identify metabolic signatures associated with malodor conditions. The investigators analyzed state-of-the art metabolomics tests to explore if conditions leading to malodor can be screened by metabolomic profiling of urine samples [40]. The study was registered on clinicaltrials.gov as NCT03582826

#### Microbiome Study

Microbial Basis of Systemic Malodor and “People Allergic To Me” Conditions (PATM) study examined gut microbiomes in different sub-cohorts and flare-ups or remission states of the studied conditions [38,58]. It was registered on clinicaltrials.gov as NCT03582826.

#### Unscheduled Visits

Unscheduled visits and inquiries will be handled and documented like scheduled visits.

### Subject Completion/Withdrawal

Subjects may withdraw from the study at any time without prejudice to their care. They may also be discontinued from the study at the discretion of the Investigator for lack of adherence to study treatment or visit schedules, or due to health reasons. The Investigator may also withdraw subjects who violate the study plan, or to protect the subject for reasons of safety or for administrative reasons. It will be documented whether or not each subject completes the clinical study. If the Investigator becomes aware of any serious, related adverse events after the subject completes or withdraws from the study, they will be recorded in the source documents and on the case report form (CRF).

#### Early Termination Study Visit

Subjects who withdraw from the study will have all procedures enumerated as the early termination visit.

## Study Evaluations and Measurements

### Screening and Monitoring Evaluations and Measurements

#### Medical Record Review

Data collection will include the following areas of interest: subject name; date of birth; home address; telephone numbers; referring physician information; alternative contacts and contact information; results of diagnostic tests.

### Safety Evaluation

There are no safety evaluations except to ensure subject confidentiality.

### Primary Endpoint

The primary endpoint is the accuracy of breath test to discriminate between individuals with systemic body malodor vs systemic breath malodor.

### Secondary Endpoints

Secondary endpoints will include the following:

- The relationship between volatile organic compounds in breath and TMAO:TMA ratio.
- The relationship between volatile organic compound composition and diet, medications, results of laboratory tests and other biomedical measurements.
- Comparison between exhaled breath VOCs of other possible sub-cohorts.

### Control of Bias and Confounding

Subjects in observational studies are not assigned by a process of randomization and are therefore subject to bias. While not completely possible to control for bias in an observational study, we have taken certain steps to minimize the risk of confounding. We will collect data on previously identified diet confounders. Biomarker measurements will be made in bulk post-clinical data collection. Thus, clinical data will be blinded to the biomarker levels or endotype assignments.

There will be no predilection towards race, ethnicity or sexual orientation. The cross-validation of the findings in this study based on other ethnicity groups and larger sample sizes will be needed for future research.

### Statistical Methods

#### Baseline Data

Baseline and demographic characteristics will be summarized by standard descriptive summaries (eg, means and standard deviations for continuous variables such as age and percentages for categorical variables such as gender).

#### Analysis of Primary Outcome of Interest

The primary analysis will include all subjects meeting all inclusion and exclusion criteria who submitted processable breath samples.

### Sample Size and Power

We established that a sample size of 10 subjects per group would be sufficient to reject the null hypothesis that VOC analysis cannot discriminate body and breath sub-cohorts with a power of 80%.

Previous studies found that 10 subjects per group could yield detectable differences in the study population [58]. It will provide 80% power if the ratio of the difference in means to the standard deviation is 1.3 (signal to noise ratio) or greater. S/N ratio of 1.5 would allow to achieve 90% power.

For Mann-Whitney test, each group’s size should be ∼11 for a P value of 0.05. For NOVA-type analyses, five subjects per group would allow 90% power to detect a ω2 (adjusted coefficient of determination) of while 10 subjects per group would allow 90% power to detect an ω2 of 0.02.

## SAFETY MANAGEMENT

### Clinical Adverse Events

Clinical adverse events (AEs) will be monitored throughout the study.

### Adverse Event Reporting

Since the study procedures are not greater than minimal risk, SAEs are not expected. If any unanticipated problems related to the research involving risks to subjects or others happen during the course of this study (including SAEs) these will be reported to the IRB in accordance with CHOP IRB SOP 408: Unanticipated Problems Involving Risks to Subjects. AEs that are not serious but that are notable and could involve risks to subjects will be summarized in narrative or other format and submitted to the IRB at the time of continuing review.

## STUDY ADMINISTRATION

### Data Collection and Management

Data will be collected as both electronic files and paper questionnaires and letters. All data will be kept secure and confidential. Paper files will be kept in a locker filing cabinet. All participants will be assigned a unique identifier for the duration of the study. A separate log will be kept linking the ID number to the participant which will be kept and stored confidentially. Patients will be identified only by their unique study ID and demographic information (age at time of study and sex) will be entered.

### Data Analysis

All data will be tested for normality. Where data are not normally distributed a nonparametric test will be performed. Data will be presented as means ± SD. Statistical analyses will be performed Statistical difference will be accepted when P < 0.05. Statistical differences will be assessed by repeated-measures.

The diagnostic accuracy of VOCs will be initially estimated by comparing its alveolar gradient values in cases and controls and determining the value of its C-statistic (“concordance” statistic, a measure of goodness of fit for binary outcomes), e.g., the AUC of the receiver operating characteristic (ROC) curve.

When analyzing breath biomarkers, we will mine the human breathomics database (HBDB) [59] general-purpose human metabolome database (HMDB) [60], microbial volatiles [61] database, and the EPA’s chemistry dashboard [62]. To map metabolic pathways, we will use the Kyoto Encyclopedia of Genes and Genomes (KEGG) database [63] and WikiPathways [64].

### Procedure for Dealing with Missing, Unused and Spurious Data

All data collected will be included in the analysis. If no data is available for a given sub-cohort analysis then the patient will be excluded from this assessment, but will still be included in other analyses. Missing data will not be imputed unless specifically noted. Any apparently spurious data will be verified. Where statistic differences are present, post hoc (Bonferroni-corrected) analysis will be performed. Outliers will remain within the data.

### Confidentiality

All data and records generated during this study will be kept confidential in accordance with Institutional policies and HIPAA on subject privacy. The Investigator and other site personnel will not use such data and records for any purpose other than conducting the study.

No identifiable data will be used for future study without first obtaining IRB approval. The investigator will obtain a data use agreement between provider and any recipient researchers before sharing a limited dataset.

## PUBLICATION

Results will be submitted for publication in a scholarly journal.

## Data Availability

available upon reasonable request

https://aurametrix.com/nct0345199.html

